# From here to there: changing the way we evaluate equitable coverage of health services

**DOI:** 10.64898/2026.07.09.26356719

**Authors:** Brittany Hagedorn, Jeremy Cooper, Anu Mishra

## Abstract

Reducing inequality in health-service coverage is central to universal health coverage, but the evidence base on how to design successful equity-oriented policies is inadequate to inform decision makers and tends to rely on case studies. Further, conventional measures of inequality capture a single timepoint, one service at a time; this obscures how inequity evolves as coverage increases. We reframe equity as a trajectory and ask how it evolves as total coverage rises, comparing systematically across countries and health areas.

Using 132 Demographic and Health Surveys from 22 low- and middle-income countries (1990–2023), we estimated coverage at the subnational (admin1) level by wealth quintile for six representative maternal and child health indicators. For each country-indicator pair, we fit a natural cubic spline of the wealthiest–poorest gap against total regional coverage, extracted features describing each curve, and grouped them using hierarchical clustering.

This yielded three archetypes: large rollout gaps (mean peak ∼58%), modest but persistent inequality (∼30%), and minimal inequality that sometimes reversed to favor the poor (∼17%). Most trajectories traced an inverted U pattern, widening early, then closing only near 100% regional coverage.

How a service is delivered, more than where, drove its path: institutional delivery was the most inequitable (15 of 20 countries with large gaps), whereas one-touch and campaign-delivered services such as bed nets and vaccines rarely produced large gaps and were sometimes pro-poor. Despite this, some countries achieved equity across nearly all services, indicating that proactive governance may be able to overcome structural challenges to achieve equitable outcomes.

For policy, these archetypes let programs anticipate which groups will be left behind and when, replace assumed scenarios with empirical ones in impact models, and target investment early to ensure that new services achieve more equitable coverage.

## Introduction

Inequality in service coverage is a primary concern for public healthcare systems around the world as governments strive to achieve universal health coverage. (1) To varying degrees, they have implemented policies that intend to reduce inequality, but the impact of these policies is rarely studied systematically across geographies and health areas. This leaves countries with good intentions but limited evidence about what works and for which services. (2) This work aims to address this by quantifying an effect size and pointing to policy and programmatic structures that have achieved better coverage equity.

When coverage is distributed unequally, this can cause two problems for public health. First is that it can create worse outcomes for everyone – e.g., through pockets of under-immunization that harbor outbreak risks. (3) Second is that it can reduce the cost-effectiveness of the intervention itself by reaching those with lower burden – e.g., prenatal iron supplements reaching those with a better diet (and thus lower anemia rates) first and reducing treatment cost-effectiveness. (4,5)

As aggregate coverage for a geography increases, there are many ways that this can evolve, either to support health outcomes and equity or to its detriment. To get from “here to there” in terms of total coverage, there are three assumed ‘curves’ that are often used for estimating potential impacts from these increases, and one more realistic scenario.

1. Everyone in the geography has **equal coverage** increases – i.e., all groups start at zero and increase at the same rate together. This rarely happens.
2. Most advantaged sub-populations see gains first (**least equitable**). For example, for costly healthcare treatments, we generally see a trickle-down in access from the wealthiest to the least as total coverage rises.
3. **Pro-equity coverage** increases, where the least advantaged groups get access first and the most advantaged only receive coverage once the others already have it. (6) This is a useful bounding scenario but rarely occurs in real implementation due to logistical, social, and political realities.
4. The most common occurrence is a **mixed inequitable curve**, where coverage across all groups is increasing over time, but more advantaged groups increase coverage faster than others. (7)

Examples of what these curves would look like are shown in Figure 1. Note that while time is not an axis on these curves, we generally observe that countries move from left to right as total coverage increases over time.

**Figure 1.**
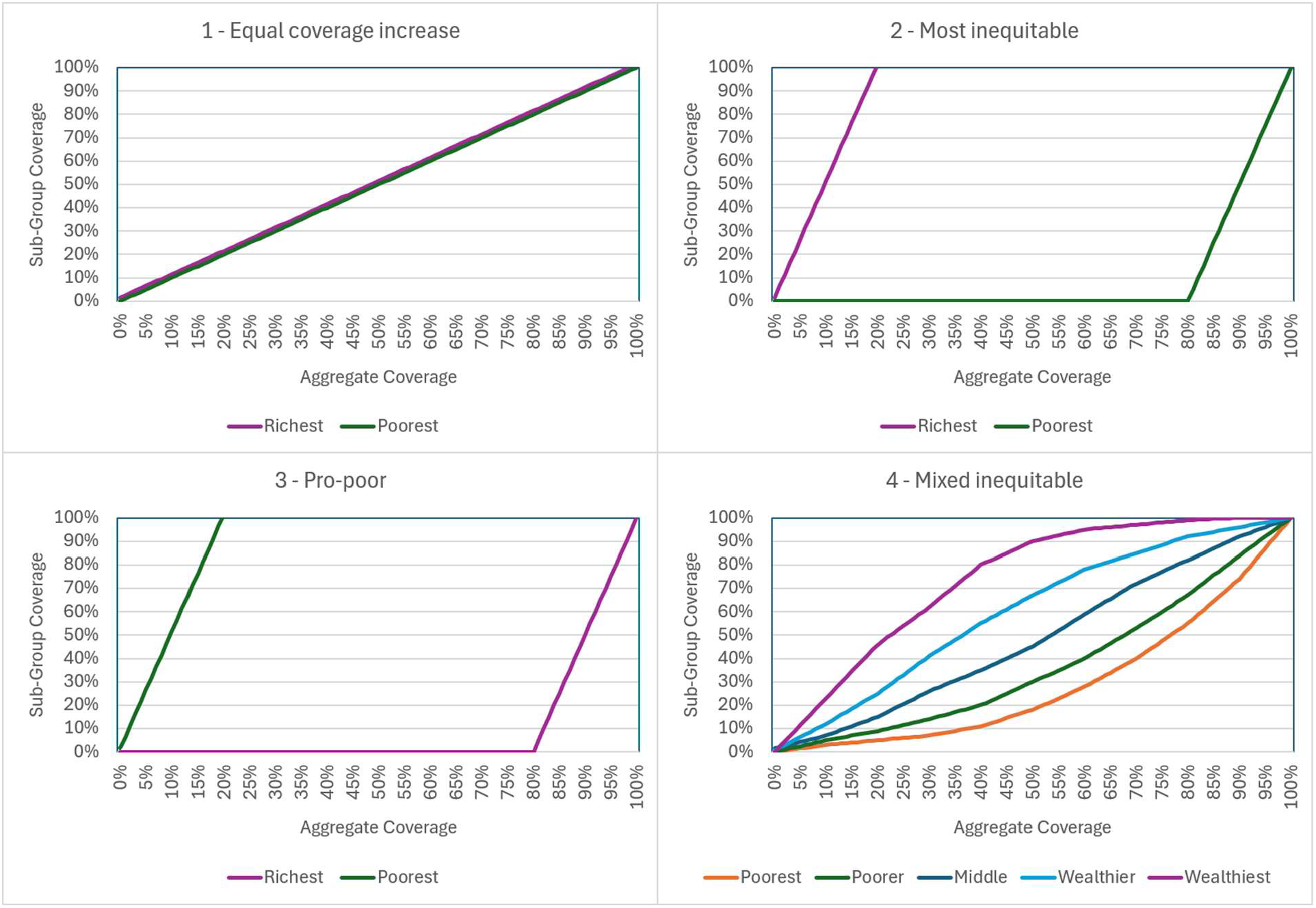
Hypothetical inequality in coverage scenario curves. Curve 1 represents equal coverage amongst all wealth groups (perfect equality). Curves 2 and 3 represent strictly minimum and maximum inequality between the wealthiest and poorest groups, respectively. Curve 4 represents more typical inequality where there is a bias toward wealthier groups getting covered first, but that is not a strict dynamic.

Current evaluation methods for inequity fall short in a couple of important ways, which this work aims to address. First, inequity is usually studied for a single service at a time (e.g., immunization, antenatal care), missing opportunities to learn across health domains.

Second, inequity is measured at a point of time, rather than tracking distributional change as aggregate coverage increases. (8,9) Note that previous work that has looked at this longitudinally has done so at the national level and with only two time points, so our work extends on this framework. (10) Third, when data is aggregated, especially to the national or regional level, it is possible to conflate increases in these total values as equity gains. Last, subnational dynamics of inequity and comparisons across regions within a country are not often studied but have been shown in limited studies to be important considerations. (11)

These differences matter for evaluation because they can have substantially different health impacts, but current methods obfuscate this. (12,13) Further, point estimates for inequality don’t tell you whether you are getting better or worse, or how to change the shape of the curve entirely. To do this, we classified the shape of inequity curve trajectories, reviewed patterns amongst groups of countries and outcomes, and assessed what this means for policies that can help countries get onto a better trajectory.

Our objective here is to better understand how coverage inequality evolves over time. Specifically, as aggregate coverage increases (e.g., nationally), to what extent and which groups are left behind, and how that varies by context.

We assessed a range of representative primary healthcare outcomes such as immunization coverage, across as many low- and middle-income countries (LMICs) as possible, to give us a large enough dataset to detect patterns and to learn lessons through comparisons across countries.

Understanding how coverage inequality evolves, not just whether it exists at a given moment, has direct implications for how health systems design and sequence their interventions. For example, a country that knows its coverage trajectory is following a common pattern where the most advantaged groups increase coverage the fastest faces a different problem than one where the least advantaged are being completely left behind. Beyond visibility into the problem, the intervention could be quite different. The former scenario may call for targeted demand-side interventions to accelerate gains among underserved groups, while the latter requires structural redesign of service delivery itself.

If policymakers can see not only where their country stands but which direction it is moving and at what rate, they are better positioned to set realistic equity targets, allocate resources to where the curve is most malleable, and hold programs accountable for distributional outcomes rather than aggregate ones alone. This paper aims to provide evidence to support that policy foundation.

## Methods

### Data and preparation

A goal of this analysis is to track measures of health equity as they change over time, at subnational levels in LMIC countries. We selected countries with at least five Demographic and Health Surveys (DHS) to ensure a minimum range of coverage for a given country-indicator pair. This resulted in 132 surveys across 22 LMICs, spanning 1990-2023.

To calculate coverage levels at the administrative1 level disaggregated by national wealth quintile, we perform a spatial join between survey clusters and GADM 410 administrative boundaries and then aggregate population weighted responses accordingly, in accordance with standard DHS practices. (14–16)

We selected six representative metrics across maternal and child health from the DHS: fourth antenatal care visit (ANC4), institutional delivery (ID), first-dose of measles-containing vaccine (MCV1), first-dose of pneumococcal-containing vaccine (PCV1), household ownership of a malaria bed net (ITTN), and care seeking rates for fever in children under age five (fever).

We defined inequality as the gap in coverage between the wealthiest and poorest quintiles, at the admin1 level, which was our primary outcome of interest. We looked at how this gap compared to the total regional (admin1) coverage, so that the result showed how inequality evolves as total coverage rises. Note that we selected this metric because it is easy to interpret and after conducting sensitivity analysis on other formulations of measuring inequality did not find a substantial difference in the results. (17)

### Four-step approach

We took a four-step approach to classify inequality for each combination of country and metric. First, we fit a spline curve to point-wise coverage estimates for each indicator across the study inclusion years, then we extracted ten features of the curves, and finally we applied clustering to the features. Last, after we identified the ‘types’ of inequity curves, we did a qualitative review of policies and contextual factors to identify the key drivers of coverage inequality.

To ensure robustness, we also performed a sensitivity analysis on indicator selection and the definition of inequality. This involved comparing alternative indicator definitions (e.g., skilled birth attendance vs. institutional delivery). We also compared alternative definitions of inequality including using the log of the ratio of the wealthiest divided by poorest coverage and considered alternative gap calculations such as combining quantile subgroups. None of these alternatives significantly affected the results, so the details are not presented here.

### Spline fitting

For each country-indicator pair, the inequity trajectory is represented as a natural cubic spline, fit to the gap between richest and poorest wealth quintiles (y-axis), as a function of total regional coverage (x-axis).

To avoid over-fitting, the spline degrees of freedom (DOF) depend on sample size. (18)

- 4 DOF: 20+ observations and 50% coverage of the 0-100% range.
- 3 DOF: 12+ observations and 40% coverage of the 0-100% range.
- 2 DOF: All others (essentially a quadratic fit).

Interior knots are placed at quantiles of the unique interior x-values, rather than at quantiles of all data. This is a deliberate departure from the default because low-coverage indicators have multiple values at 0% coverage, which would collapse the default knot placement onto the boundary.

Anchor points at (0, 0) and (1, 0) are appended to enforce the true mathematical constraint that no gap can exist when total coverage is 0% or 100%. This also prevents unconstrained tails from dominating the fitted form. To reduce noise in the final analysis, we only fit splines to datasets with at least ten datapoints, at least 5 distinct x-values, and a range of values covering at least one third of the 0-1 potential x-range.

### Curve features

We extracted eleven features for use in the clustering analysis. The features include items to assess the level of inequality at its peak, the degree to which inequality is high or low across the curve, and the shape of the curve including skewness. This results in the following features, which are described in detail in the supplement.

- Location of the spline peak (x and y values),
- Number of local spline maxima (1+),
- Portion of the 0-100% range that is covered by the data (x range),
- Mean squared error, comparing the spline fit to the input data,
- Area under the spline curve (AUC),
- Portion of spline where the curve that is ‘high’ (above 80% of the peak value),
- Asymmetry ratio, comparing the portion of the spline curve that precedes vs. follows the band where the curve is high,
- Extent and location of sub-zero curve segment (i.e., where the poorest quantile had more coverage than wealthiest).

### Clustering

We utilize hierarchical clustering as a partitioning method, as implemented in the hclust() function in R. This uses Ward’s minimum-variance linkage, which minimizes the within-cluster sum of squared deviations at each merge and aligns with the goal of identifying compact, distinguishable trajectory archetypes. (19)

To evaluate clustering fit, we review cophenetic correlation, an explicit “merge-jump” analysis that ranks the cluster counts from largest to smallest (large = better), within-cluster sum of squares elbow plot, and silhouette widths. (20,21)

## Results

### Dataset

We identified 22 LMICs with at least five separate DHS datasets, which resulted in a total of 138 potential country-metric pairs. After feature extraction and checking sample size, we rejected 19 country-indicator pairs because they did not have data covering at least 33% of the total coverage range and two pairs because they did not have adequate sample size. This resulted in a net of 117 country-indicator pairs for which we fit spline curves and extracted features.

After reviewing the cluster fit metrics, we chose k=3 as the best-fit number of clusters. (See supplement for the detailed cluster metrics on which we made this decision.) We then visualized the mean and standard deviation of the features that were used to create the clusters and used this to name them in a rational, human-readable way. These are summarized in Figure 2.

**Figure 2.**
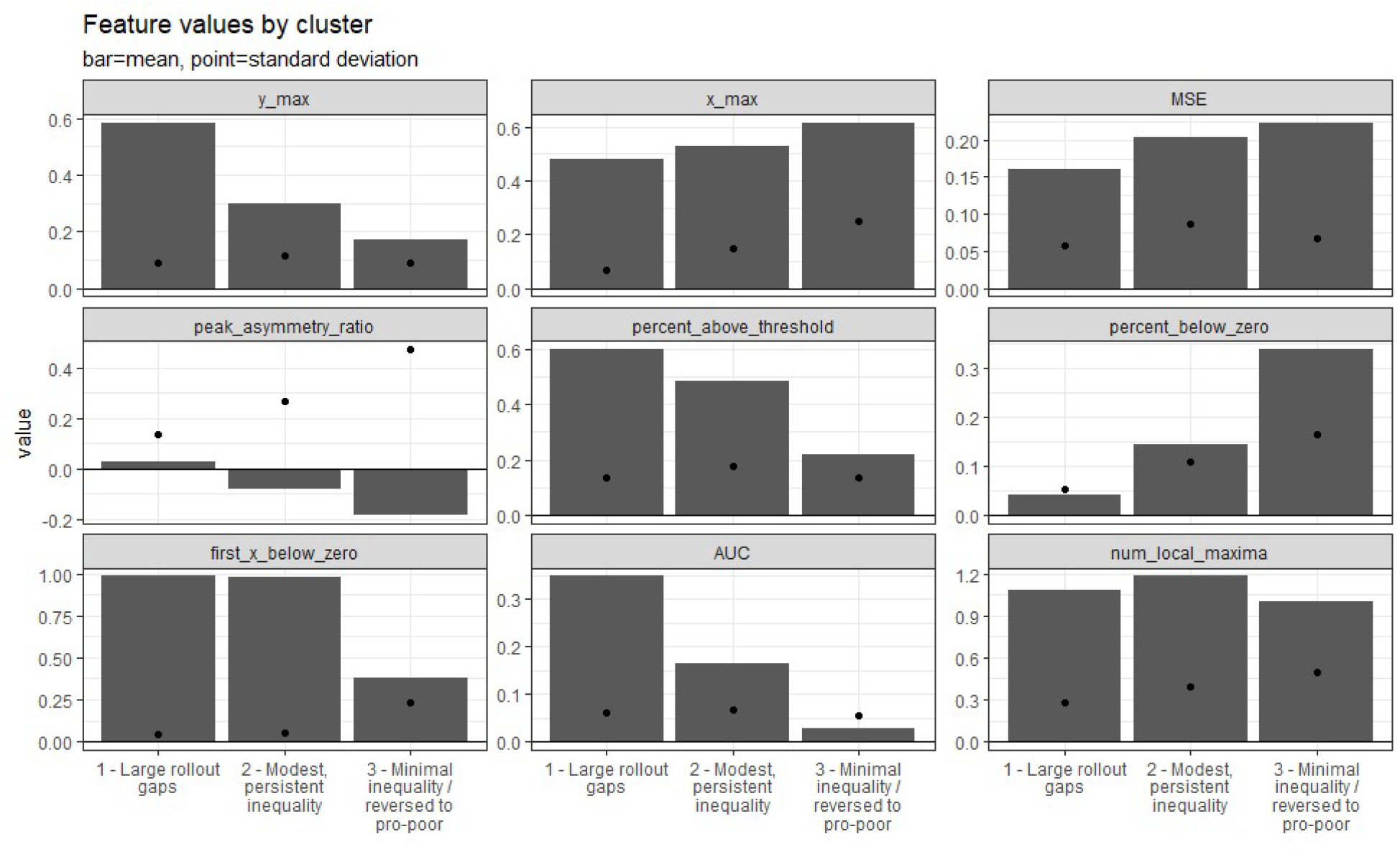
Values of the features extracted from fitted spline curves, by cluster. Bars represent mean values for all curves in the cluster and points are the standard deviation. Detailed definitions of the metrics are found in the supplement. Cluster names were written to summarize these key characteristics.

In order to verify that the clusters correspond to distinct curve trajectories and uncover patterns, we examined the fitted splines. This is shown in Figure 3. Individual country-metric cluster assignments are shown in the supplement.

**Figure 3.**
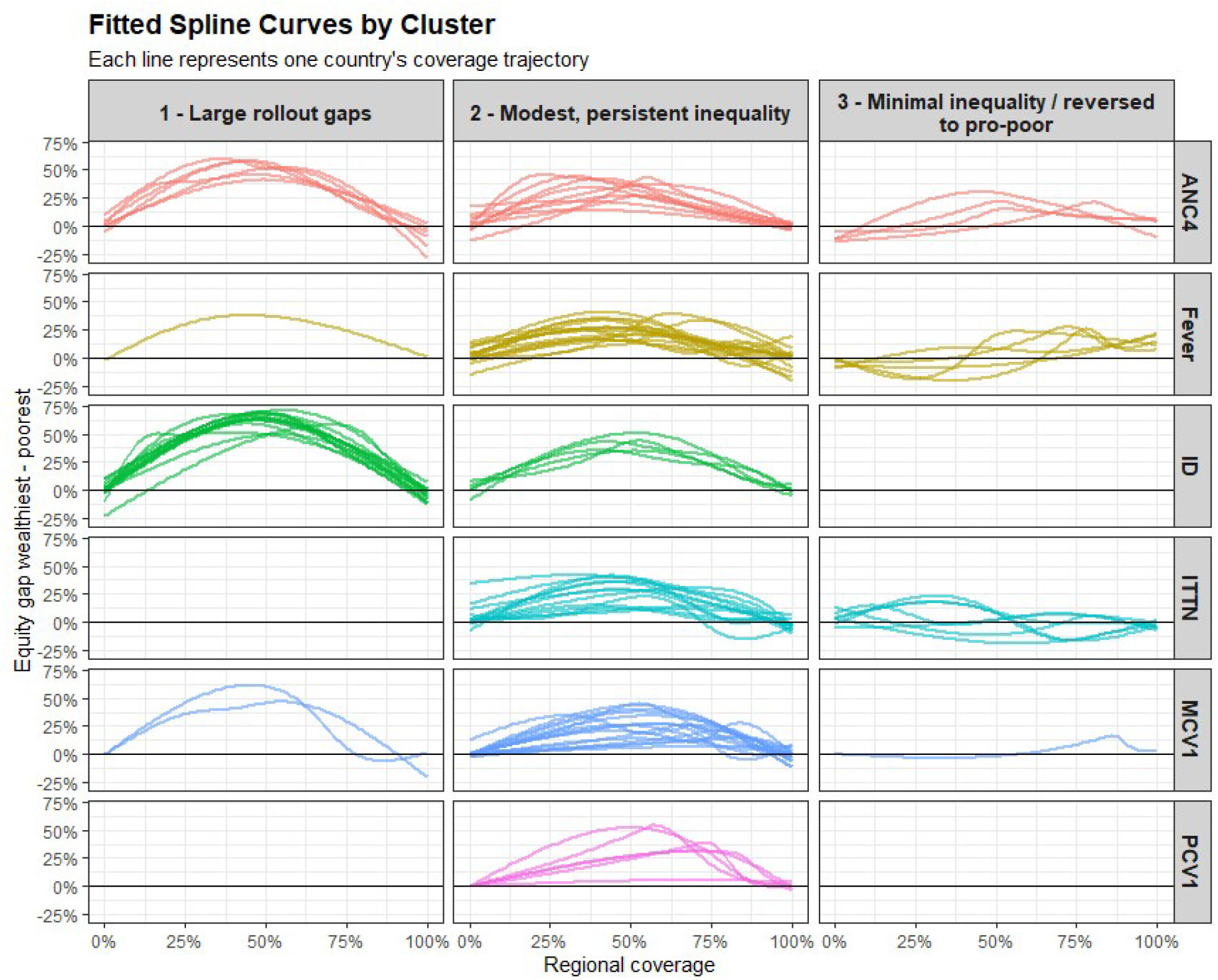
Spline fits, split by cluster and outcome metric. Each curve represents a different country’s data, with regional coverage shown on the x-axis and the equity gap (as defined by the gap in coverage between the wealthiest and poorest quintiles) on the y-axis. Higher, broader curves above the 0% line indicate pro-wealthy inequity. Curves near the 0% line indicate equal coverage in the wealthiest and poorest quintiles. Curves that dip below 0% represent instances when coverage was pro-poor.

Here we see that services have different clustering patterns, with some being distributed more equitably than others. Institutional delivery (ID) has the highest overall inequality, with 15 of 20 countries seeing large rollout gaps and zero countries falling into the minimal inequality cluster. Antenatal care (ANC4) has the most mixed profile, with six countries having large gaps, ten having modest persistent inequality, and four minimal inequality. The remaining outcomes of care seeking for fever, ITTNs being available in households, and the vaccines MCV1 and PCV1 are mostly centered in the modest inequality cluster.

The cluster where we observed minimal inequality and sometime reversed, pro-poor coverage gaps was composed of seventeen country-metric pairs. These included:

- ANC4: Kenya, Lebanon, Malawi, and Uganda
- Fever: Lebanon, Malawi, Northern Nigeria, Zimbabwe, and Zambia
- ID: (none)
- ITTN: Guinea, Ghana, Cote d’Ivoire, Moldova, Northern Nigeria, Southern Nigeria
- MCV1: Tanzania
- PCV1: (none)

Finally, when we look at individual countries, we see that some have a more pro-equity set of curves than others. (See Figure 4.) Of the countries that were evaluated on at least five outcome metrics, eight had no outcomes with large rollout gaps, including the sub-Saharan countries Benin, Burkina Faso, Malawi, Rwanda, and Uganda. Of note, Malawi and Uganda also had at least one metric classified as ‘minimal inequality’, suggesting that they may had pro-equity strategies in place. An additional six countries had only one metric with a large gap: Ghana, Kenya, Mali, Mozambique, Senegal, and Tanzania. At the other end of the spectrum, the two countries most frequently classified into the ‘large gap’ cluster were Ethiopia (3 of 5 metrics) and Nigeria (3 of 6 metrics).

**Figure 4.**
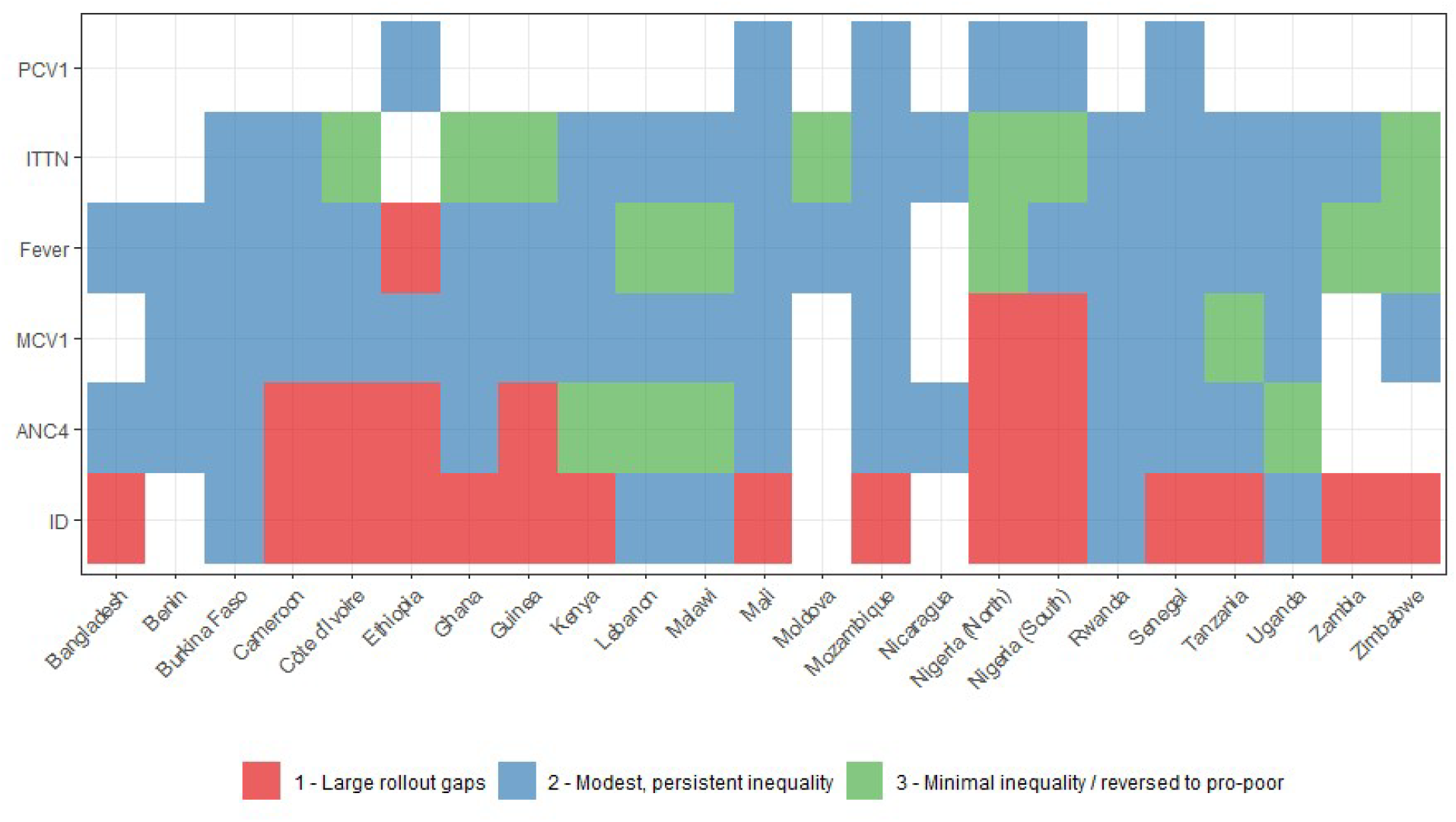
Curve classifications into each cluster, by metric and country.

## Discussion

### Principal findings: inequality as a trajectory, not a point

Across services and countries, inequality in service coverage behaved less like a fixed attribute of a place or program and more like a predictable, transient feature of coverage scale-up. When the gap between the richest and poorest wealth quintiles is plotted against regional coverage, most trajectories traced an inverted U with a widening gap as wealthier groups gain access first, consistent with previous findings of “inverse equity”. (7) The constraint that no equity gap can exist at 0% or 100% coverage is mathematical, but the height, position, and persistence of the hump in between are not – and these features separate a system that briefly leaves the poor behind during early rollout from one with sustained inequality.

Understanding this requires us to reframe equity as a trajectory, not a static feature of a particular service. This matters because a point-in-time inequality statistic cannot tell a policymaker whether a country is headed in the right direction or if a policy change is needed; the trajectory can.

Clustering allowed us to identify three interpretable archetypes: large rollout gaps, modest but persistent inequality, and minimal inequality that sometimes reverses to favor the poor. These differ chiefly in the peak magnitude of the gap (cluster-mean peaks of 58%, 30% and 17%, respectively), in how long the curve remains high near the peak, and in whether and where it crosses into pro-poor territory. The minimal-inequality cluster was particularly distinguished by pro-poor gaps in coverage and by a front-skewed curve, consistent with inequity that appears during early rollout but then dissipates rather than entrenching.

### Service delivery is the primary driver

The strongest signal in the results is that *how* a service is delivered, more than *where*, drives its equity trajectory. Institutional delivery was the most inequitable service by a wide margin: 15 of 20 countries fell into the ‘large rollout gap’ cluster and none were classified as ‘minimal inequality’. This is consistent with institutional delivery being the most demanding service in our set. It requires costly physical infrastructure and skilled staff, is effective only in a narrow time window, and competes with alternatives such as traditional birth attendants. Each of these barriers falls hardest on the poorest and most remote households, so the wide inequity gap opens early and is hard to resolve. (7)

At the other extreme, services delivered through national vertical programs that only require a single contact (ITTNs, MCV1, and PCV1) almost never produced large equity gaps. In particular, ITTNs frequently exhibited pro-poor dynamics, the signature of mass distribution campaigns designed to reach those most at risk.

The clearest illustration is within Nigeria. In both northern and southern areas, facility-based and routine services (institutional delivery and MCV1) fell into the large-gap cluster, while bed nets and care-seeking for fever had minimal inequality. The fact that the country can deliver one service in a pro-poor way and another pro-rich points squarely at delivery modality and governance, rather than socioeconomic context, as the key driver.

### Proactive governance can overcome structural barriers

Despite the importance of delivery mode, it is not the whole story. The same outcome metric fell into different clusters across countries, and at the national level some systems achieved broadly equitable profiles across nearly every service they offered. Of the countries evaluated on at least five metrics, eight had no service in the large-gap cluster, including Sub-Saharan countries Benin, Burkina Faso, Malawi, Rwanda, and Uganda. Further, Malawi and Uganda achieved the minimal-inequality cluster on multiple metrics, a pattern that is difficult to explain without some intentional, system-wide orientation toward equity. At the opposite end, Ethiopia and Nigeria both had at least three outcome metrics in the large-gap cluster.

The coexistence of a strong delivery effect with a residual country factor suggests that equitable coverage is achievable but not automatic. Favorable delivery channels make it easier, while deliberate national prioritization appears able to extend equity even into the services that are structurally hardest to deliver fairly.

### Implications for product introductions

Comparing services at different stages of their lifecycle provides additional nuance. MCV1, long established within routine immunization, was distributed across all three clusters, whereas the more recently introduced PCV1 clustered in the modest-but-persistent inequity group. We conclude that PCV1, by riding on the established immunization system, inherited enough of that platform’s reach to avoid the large rollout gaps of older programs, but has not yet had the time to resolve inequitable access gaps.

A parallel contrast between routine preventive services (MCV1, PCV1, and ANC4) and acute care (fever care-seeking and institutional delivery) follows the same logic: preventative services equalize more readily than those that depend on a household seeking care promptly.

Looking forward to other new products, the equity trajectory is likely predictable based on the maturity and delivery modality of the platform onto which it is introduced. This makes a strong case for continued investment in existing touch points such as antenatal care, as a platform for future interventions to be introduced.

### Methodological contribution and implications for modeling

Methodologically, this work demonstrates the value of moving from point estimates to the classification of inequality *trajectories*, and of doing so at subnational resolution. Pooling surveys into a common, longitudinal dataset is what turned idiosyncratic country narratives into comparable archetypes, providing deeper insight than any single survey, indicator, or point estimate could have yielded on its own.

Technically, an empirically based trajectory should replace the stylized “equal,” “pro-rich,” and “pro-poor” scenario curves that modelers currently assume when projecting the impact of coverage scale-up; rather than bounding an unknown, a model can anchor to the trajectory archetype that a given service and context most resemble.

Programmatically, knowing in advance which groups a given delivery modality tends to leave behind, and at what point along the coverage range the gap is widest, allows mitigation to be designed before a product is introduced or a scale-up target is set. This gives decision makers who are trying to achieve equity goals the opportunity to intervene in advance, rather than diagnosing the problem after inequity has already emerged. Because the curve is most malleable on its leading edge, this also indicates where additional investment is likely to buy the most equity.

### Limitations

Several limitations temper these conclusions. Our equity measure captures the economic dimension of inequality but not geographic, gender, or ethnic disparities, nor inequality within quintiles; sensitivity analyses using ratio-based and combined-quintile definitions did not materially change the findings, but the wealth axis remains only one lens. The trajectories are constructed across coverage levels observed in different regions and survey rounds rather than by following a single region through time, so they are best read as a space-for-time approximation of how inequality tends to evolve rather than as a path any one region is guaranteed to take. The links between clusters and their hypothesized drivers – delivery modality, funding source, program maturity, and national prioritization – are qualitative and associative; this design cannot establish that any policy caused a given trajectory. Finally, although the three-cluster solution was clearly favored by the merge-distance criterion, it rests on modest silhouette widths, so the archetypes are best treated as useful summaries of an underlying continuum rather than as sharp, natural categories.

## Conclusion

Taken together, these results recast coverage inequality as a dynamic and largely predictable phenomenon, rather than a static problem. The dominant shape – inequality that emerges during rollout and resolves at saturation – means the policy-relevant questions are how high the gap rises, how long it lasts, and whether it ever reverses, all of which our trajectory archetypes capture and a point estimate cannot. How a service is delivered emerges as the primary lever: facility-based, time-critical care such as institutional delivery is the hardest to equalize, whereas campaign-style, one-touch, often donor-funded interventions reach the poorest most readily and sometimes first. Yet country experience shows that even structurally difficult services can be delivered equitably where systems make it a priority, demonstrating that pro-equity coverage is an achievable design goal rather than merely an aspiration. By making the trajectory visible, this approach offers programs a way to anticipate who will be left behind and when, to replace assumed scenarios with expected ones, and to act while the curve is still malleable, turning the measurement of inequality from a retrospective verdict into a forward-looking instrument for getting from here to there more equitably.

## Data Availability

No new data was produced in this study. All data used for the analysis was obtained through the Demographic and Health Survey (DHS), which is available online upon request.

## Supplement

### Feature Extraction Summary

**Mean Squared Error** is the residual standard error of the fit, evaluated only on interior data points; a prior version of this calculation compared predicted y to x through partial-name matching and silently returned near-zero values, so the current code uses an explicit residual sum of squares against the true gap between richest and poorest.

The location and magnitude of maximum inequity are summarized by **the y peak max** (the peak value of the smoothed curve over observed support) and the **x location of the peak** (the coverage level at which that peak occurs). Peaks are calculated from the spline, which was built on the observed data (not the full 0-100% potential range), so that extrapolated tails outside the country’s experience cannot generate phantom peaks.

Three features describe the shape of the inequity trajectory.

The **percentage above threshold** is the fraction of observed support over which the curve sits above 80% of its peak value; it captures how long inequity persists near its maximum, distinguishing sharp transient gaps from broad plateaus of persistent inequity. This defines the ‘band’ of high inequality used in other feature calculations.

The **peak asymmetry ratio** complements this by locating that above-80% band on the shared [0, 1] coverage axis. It is defined as (1-portion above band) – portion below band, taking values in [−1, 1]. Positive values indicate the inequity band sits early in the coverage spectrum (early-rollout inequity that resolves), values near zero indicate symmetry around 50% coverage, and negative values indicate that high inequity persists into high-coverage settings (late-rollout persistence). Although the band itself is identified on observed data only, the asymmetry is measured against the universal 0 and 1 endpoints so that countries with different supports remain comparable.

The **number of local maxima** counts local maxima of the curve (with the 0 and 1 anchors included, since they are real constraints on the spline). This value flags non-monotone shapes such as double-humped trajectories.

Two features describe the extent to which the data reveals any area of the distribution that is pro-equity (i.e., where poorest have higher coverage than wealthiest).

The **percentage below zero** is the fraction of admin1 observations for which the richest-minus-poorest gap is already negative, i.e., where the poorest quintile has higher coverage than the richest. This may reflect targeted programs or saturation effects.

The same phenomenon is captured on the spline curve by calculating the **first x-value where the spline crosses zero** and the gap turns negative. When the curve never crosses zero within the observed data range, this is set to 1. If the curve dips below zero, rises above, and dips again (i.e., there is a non-contiguous negative region), we select the largest region and use that crossover value, so that the feature represents the durable pro-equity region rather than a brief wobble near the axis.

One feature summarizes the total magnitude of inequity.

**Area under the curve** (AUC) is the mean value of the spline, evaluated on the full 0 to 100% coverage region. We evaluate across the full range so that countries are integrated over a common domain and so the anchor-tied tails dampen the curve where the country has limited data. This measures the average intensity of inequity. Note that it is possible for pro-wealthy inequality to be ‘ofset’ by the inverse pro-poor inequality in another part of the curve.

### Clustering Results

#### Merge Distances

- k = 3 (merge distance jump: 6.17)
- k = 6 (merge distance jump: 2.71)
- k = 4 (merge distance jump: 2.06)
- k = 5 (merge distance jump: 1.81)
- k = 13 (merge distance jump: 1.12)

#### Elbow Method

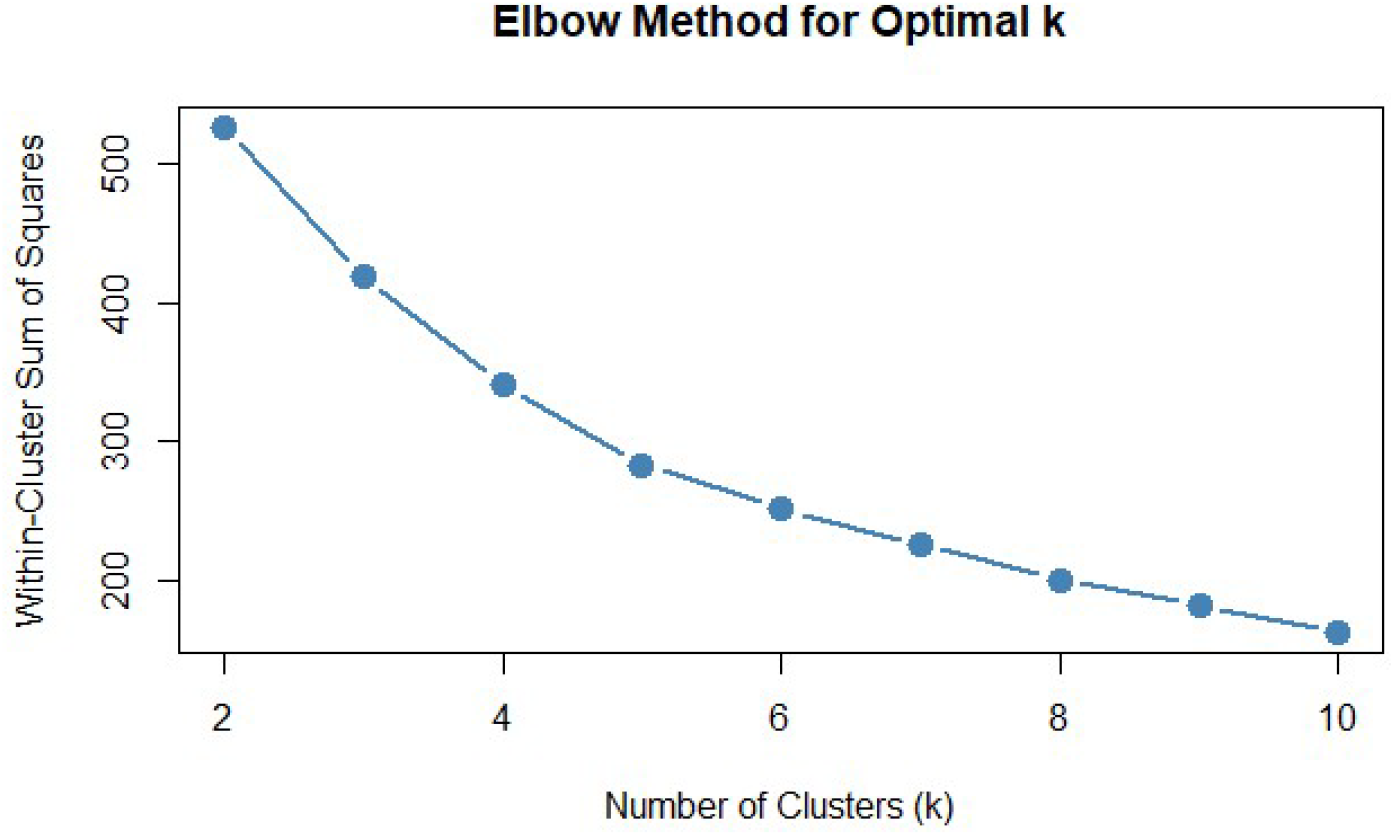

#### Silhouette Width

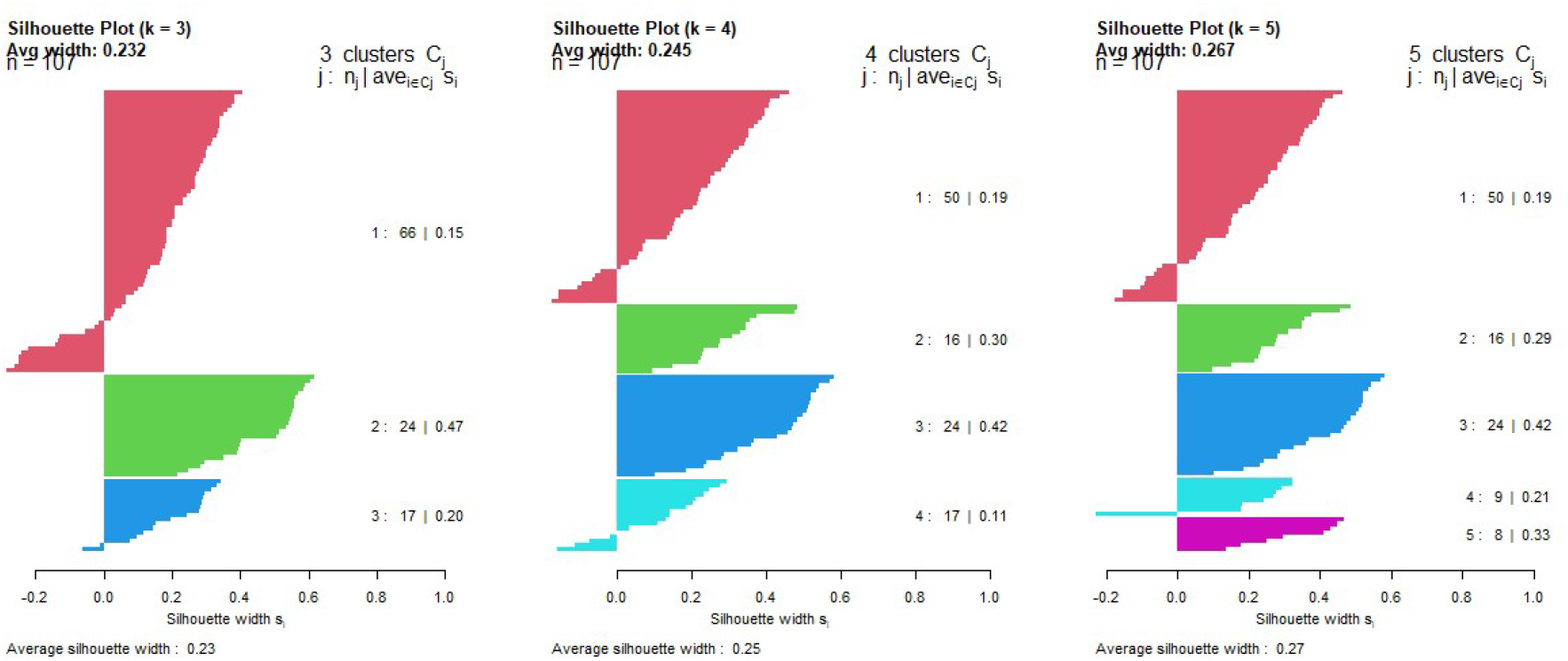

